# Powassan Virus Seroprevalence in a U.S. Servicemember Population at High Risk for Tick Exposure

**DOI:** 10.64898/2026.06.02.26354611

**Authors:** Alexandra L. Tse, Zoey Dipasqua, Joelle El Hamouche, Georgia Fallon, Kira E. Enos, Griffin C. Horowicz, Michael J. Rossen, Wyatt V. Chapman, McKenzie N. Daffin, Kayla A. Kiniry, Alexis Jankovich, Joshua S. Choy, Audrey R. Whitfield, Beth A. Bachert, Erik Cazares, Gorka Lasso, Justin E. Jones, Stacey L. Bateman, Daniel Gordon, Shauna L. Stahlman, Andrew S. Herbert, Catalina Florez, Jonathan R. Lai, Kartik Chandran, Kevin J. O’Donovan, Jeremy R. Hershfield, Emily Happy Miller

## Abstract

Powassan virus (POWV) is an emerging tick-borne flavivirus that can cause severe encephalitis in humans. Currently no vaccines or therapeutics are approved to treat POWV. POWV is spread by the deer tick, *Ixodes scapularis*, which is ubiquitous across the Northeastern United States. To better understand POWV prevalence in high-risk populations, we examined POWV seroprevalence in Cadets at United States Military Academy (USMA) in West Point, New York. Cadets at USMA, located in a heavily wooded area, are at high risk for tick exposure during outdoor military training. 1,051 serum samples from the Cadet class of 2017 were screened for POWV seropositivity using a POWV Envelope (E) DIII ELISA. A seropositivity rate of 1.3% was determined. Several ELISA-positive samples were also able to neutralize both reporter virus particles bearing the POWV E protein and authentic POWV. This study demonstrates populations at risk for tick exposure may have significant seroprevalence of POWV.

## Background

Powassan virus (POWV) is an emerging tick-borne flavivirus that can cause severe encephalitis and meningitis in humans. Approximately 10-15% of neuroinvasive POWV infections are fatal, and among survivors, nearly half experience long-term neurological sequelae. Currently, there are no approved vaccines or therapies available for POWV infection. Although the overall incidence remains low, reported cases in the United States have risen in recent decades^1, 2^, with 270 cases documented between 2014 and 2023 - a fourfold increase compared to the previous decade^3^. These numbers likely underestimate the true burden of disease, as asymptomatic and mild infections are rarely recognized or tested. Estimates suggest that 3,000-5,000 POWV transmission events from tick exposures may occur annually in the United States. Notably, POWV transmission from the vector *Ixodes scapularis* is rapid, with virus transfer occurring in as little as 15 minutes^4, 5^. Prior studies examining the prevalence of POWV in *Ixodes scapularis* ticks within states that have had reported human POWV cases have ranged from <1% - 5%^6–8^. With expanding tick ranges and lengthening of the tick season associated with shifting climate dynamics, POWV infections are likely to continue increasing.

POWV is an enveloped positive-strand RNA flavivirus whose envelope (E) glycoprotein mediates viral entry. The E protein consists of three domains (EDI, EDII, and EDIII)^9^ and adopts a head-to-tail prefusion E dimer configuration that is characteristic and highly conserved among flaviviruses^10, 11 12^. EDI functions as a molecular hinge linking membrane-proximal EDIII with the fusion loop-containing EDII^10, 11^. EDIII is an immunoglobulin-like domain implicated in receptor attachment in several flaviviruses and a major target of neutralizing antibodies^9^. Consistent with findings in Dengue virus (DENV) and West Nile virus (WNV), monoclonal antibodies targeting POWV EDIII demonstrate neutralizing and protective activity^13–15^. In mice, EDIII-based immunization elicits a robust neutralizing and protective antibody response^15^, and EDIII has been previously used to screen for POWV seropositivity^16^. Antibodies against POWV EDIII serve as an important basis for both serological screening assays and therapeutic antibody development.

To better understand the prevalence of non-neuroinvasive POWV infection, we developed a POWV EDIII-based ELISA screen to examine seroprevalence in a retrospective cohort from the Corps of Cadets at the United States Military Academy (USMA) located in West Point, New York. This population is at high risk for tick-borne infections due to year-round field training in wooded areas of the lower Hudson Valley, where POWV-positive ticks have been identified^7, 17^. In this study, we evaluate POWV seroprevalence in a large cohort (n=1051), providing insight into the risk of non-neuroinvasive POWV disease in a high-risk population.

## Methods

### Sample Collection and Sera Preparation

Specimens from the Department of Defense Serum Repository (DoDSR) were obtained for Cadets who entered USMA in summer 2013 (Class of 2017)^18^. From this roster, the Armed Forces Health Surveillance Division (AFHSD) extracted two specimens for each service member: 1) serum collected closest to June 30, 2013 (the “Pre” specimen), and 2) serum collected closest to June 30, 2017 (“Post” specimen). Specimens were labeled with a unique ID number and de-identified data were provided to the research team. Individuals were included in the overall study population if either a “Pre” or “Post” specimen was available. Prior to use in assays, specimens were thawed on ice and heat-inactivated at 56°C for 30 minutes to deactivate complement. This study met the requirements of the Institutional Review Board’s exempt status by the USMA Human Research Protections Program (Exempt Approval Project Number: 18-126).

### Recombinant POWV EDIII Proteins

#### POWV EDIII-MBP

POWV EDIII was produced as a fusion to maltose-binding protein (MBP) as previously described^15^. Plasmid encoding POWV EDIII-MBP was transformed in *E. coli* BL21(DE3) cells (NEB), and protein expression was induced at OD_600_ of 0.6–0.8 with 0.4 mM Isopropyl-β-D-thiogalactopyranoside (IPTG) for 18 hours at 22°C. Cells were lysed by sonication in 50 mM Tris pH 8.0, 250 mM NaCl, and MBP-POWV fusion proteins were purified by Nickel-NTA chromatography and confirmed by SDS-PAGE.

#### POWV EDIII

Gene fragments encoding the soluble codon-optimized POWV-II EDIII with a C-terminal deca-histidine and Avi tag were obtained from Twist Biosciences and cloned into pCAGGS mammalian expression vector. Plasmid sequences were verified by Sanger Sequencing. Proteins were expressed in Freestyle 293-F (ThermoFisher Scientific, R79007) cells using linear polyethylenimine (PEI, Polysciences) transfection according to the manufacturer’s protocols. Supernatants were collected six days post-transfection and proteins were purified by Ni-NTA affinity chromatography (Qiagen). Proteins were stored in 150mM HEPES and 200mM NaCl (pH 7.4) at −20°C.

### POWV EDIII binding ELISA

96-well half-area high-binding plates (Corning) were coated overnight at 4^°^C with either 20 ng POWV EDIII-MBP or 125 ng POWV EDIII. Plates were washed with 1x phosphate-buffered saline (PBS) containing 0.05% Tween (PBS-T) solution and blocked with 5% non-fat milk in PBS (blocking buffer). Heat-inactivated sera were added in triplicates as two-fold serial dilutions starting at 1:30 and incubated for 1 hour at 37°C. The POWV mouse monoclonal antibody, m158.25 was added to POWV negative sera at 26.7nM as a positive control in ELISA binding assays^15^. Plates were washed with PBS-T and stained with goat anti-Human IgG HRP secondary antibody (Rockland Immunochemicals). Plates were washed with PBS-T, TMB Liquid Substrate (Sigma-Aldrich) was added, and plates were developed in the dark for 5 minutes. An equal volume of 0.5M sulfuric acid was added to stop the reaction, and absorbances were read at 450 nm on a Cytation 5 cell imaging multi-mode reader or Synergy 4 microplate reader (BioTek Instruments).

### Borrelia burgdorferi ELISA

Selected samples were evaluated for *Borrelia burgdorferi* IgG antibodies using the SERION ELISA classic *B. burgdorferi* IgG kit (ESR121G; SERION Immunologics). Serum was diluted 1:100 in manufacturer-provided *B. burgdorferi* IgG/IgM sample buffer containing *Treponema phagedenis* lysate to reduce cross-reactivity from non-Borrelia spirochete antibodies. and rheumatoid factor absorbent was added to minimize nonspecific IgM interference. Diluted samples were added to manufacturer provided ELISA microtiter plates coated with *B. burgdorferi* antigen and incubated for 60 minutes at 37°C, followed by washing with manufacturer provided wash buffer. Plates were incubated with polyclonal goat anti-human alkaline phosphatase antibody for 30 minutes at 37 °C, washed, and developed with para-nitrophenylphosphate, solvent free buffer preservative: <0.1% sodium azide for 30-minutes at 37°C. The reaction was stopped with 1.2 N sodium hydroxide and absorbance was measured at 405nm on a Synergy 4 plate reader (BioTek Instruments).

### POWV reporter virus particle (RVP) generation

Open reading frames encoding POWV-II (GenBank: HM440558.1) structural proteins (C, prM, and E) were codon-optimized for expression in human cells, synthesized, and cloned into the pCAGGS expression vector by Epoch Life Science. The WNV subgenomic replicon-expressing plasmid (pcDNA6.2-WNIIrep-GFP/zeo) was kindly provided by T. Pierson. For RVP production, 293FT cells were co-transfected with WNV replicon and C-prM-E expression plasmids at a 1:3 ratio. Eight hours post-transfection, culture medium was replaced with low-glucose DMEM (Life Technologies) supplemented with 5% fetal bovine serum and 25mM HEPES. Cells were then incubated at 37°C with 5% CO_2_ for 3-4 days. Supernatants were harvested, clarified, and RVPs were pelleted by ultracentrifugation through a 30% (v/v) D-sucrose cushion using a SW28 rotor in an Optima LE-80K ultracentrifuge (Beckman Coulter) at 28,000 rpm for 4 hours at 4°C. RVPs were resuspended overnight on ice in NT buffer (10 mM Tris, pH 7.5, 135 mM NaCl), aliquoted, and stored at −80°C.

### POWV RVP binding ELISA

High-binding half-area 96-well plates (Corning) were coated with 25ng 4G2 mAb overnight at 4°C^19^. Plates were washed with PBS and incubated with pre-titrated POWV-II RVPs for 1.5 hours at 37°C. Plates were washed with PBS and blocked with blocking buffer for 1.5 hours at 37°C. Blocked plates were washed with PBS and exposed to a 3-fold dilution series of heat-inactivated sera or control mAb for 1.5 hours at 37°C. After washing, binding was detected with goat anti-human IgG HRP (Invitrogen) for 1 hour at 37°C. Plates were washed, and 1-Step Ultra TMB-ELISA Substrate Solution (ThermoFisher) was added and quenched with sulfuric acid. Absorbances were read at 450 nm on a Cytation 5 cell imaging multi-mode reader (BioTek Instruments).

### POWV RVP Neutralization Assay

Heat-inactivated sera or mAb were serially diluted in DMEM (ThermoFisher), supplemented with 2% fetal bovine serum (heat-inactivated, GeminiBio), 1% Glutamax (ThermoFisher), and 1% penicillin-streptomycin (ThermoFisher), and incubated at room temperature with POWV-II RVPs for 1 hour. Sera or mAb-virus mixtures were added in triplicates to Vero cell (American Type Culture Collection (ATCC), CCL-81) monolayers pre-seeded in 96-well plates (Costar) and incubated for 24 hours at 37°C and 5% CO_2_. Cells were fixed with 4% paraformaldehyde (Sigma-Aldrich), washed with PBS, and detected using a monoclonal anti-GFP antibody (Sigma-Aldrich) and an Alexa Fluor 488-conjugated goat anti-mouse secondary antibody (ThermoFisher). Infection was quantified using a Cytation-5 automated fluorescence microscope (BioTek) and analyzed with Gen5 data analysis software.

### Authentic POWV Neutralization Assay

POWV II (Spooner) was provided by the University of Texas Medical Branch (UTMB) Arbovirus Reference Collection. A low passage viral stock was generated by infecting Vero E6 cells and harvesting the cell supernatant 5 days post-infection. Supernatant was clarified by centrifugation at 2,000 xG (Beckman Coulter Avanti J-E; #369001) for 10 minutes. Viral titers were determined by plaque assay using Vero E6 cells. For microneutralization assays, pre-titrated amounts of virus were mixed with serially diluted serum for 1 hour before adding virus/serum mixture to A549 cells at an MOI of 0.02. Following 48 hours incubation, viral infectivity was determined by immunostaining formaldehyde-fixed and permeabilized cells with a mouse anti-NS1 monoclonal antibody (Native Antigen; M155) diluted 1:500. A secondary goat anti-mouse antibody conjugated with Alexa Fluor 488 (ThermoFisher Scientific) was used to detect infected cells. Images were acquired on a Cytation 5 cell imaging multi-mode reader (BioTek Instruments) and enumerated using the system’s onboard Gen5 software.

### WNV EDII binding ELISA

Sera were analyzed for reactivity to WNV EDIII using the ELISA assay described above, with the following modifications: 96-well half-area high-binding plates (Corning) were coated with 125 ng WNV EDIII (Sino Biological) at 4°C overnight. The humanized WNV mouse monoclonal antibody E16 (provided by L.M Walker) was added to WNV negative sera at 65 nM for a positive control^14^.

## Results

### POWV seropositivity in the USMA 2017 Corps of Cadets

POWV seroprevalence was determined in serum samples collected closest to graduation (“Post”) from 1,051 cadets from the USMA Class of 2017 (Table 1). The sex composition was 84.3% male (n=874) and 15.7% female (n=163). The race composition was 71.2% white (n=738), 8.0% black (n=83), 9.0% other (n=93), and 11.9% unknown (n=123). For “Post” serum samples, Cadets were between 21 and 28 years old, with the average age approximately 23.0 years. 14/1,051 samples lacked demographic information. 989 of the 1051 serum samples had known home of record data. 968 of the 989 were from the US or US territories. The participants originated from all 50 states and 4 territories in the United States (Supplementary Table 1). The northern Midwest demonstrated the lowest number of study participants (47). Approximately 290 (27.5%) cadets were from states with reported POWV cases^3^.

**TABLE 1.** DEMOGRAPHICS OF STUDY POPULATION.

|  | *“Post” Samples<sup>1</sup>* |
| --- | --- |
| Gender, n (%)<sup>2</sup> |  |
| Male | 874 (84.3%) |
| Female | 163 (15.7%) |
| Race, n (%) |  |
| White | 738 (71.2%) |
| Black | 83 (8.0%) |
| Other | 93 (9.0%) |
| Unknown | 123 (11.9%) |
| Age, mean (SD) | 23.0 (1.16) |
| Minimum | 21 |
| Maximum | 28 |

<sup>1</sup>“Post” samples from the USMA Corps of Cadets Class of 2017 correspond to those collected closest to June 30, 2017
<sup>2</sup>Percentages are based on 1,037 samples. 14 additional samples lacked demographic data. SD, standard deviation

To determine whether cadets may have been exposed to POWV, “Post” serum samples were screened for reactivity using a POWV EDIII ELISA. In the initial screen, POWV EDIII fused to MBP was utilized as antigenic bait (Figure 1A). Any sample with an absorbance at 450nm above the mean negative control sera was included in the second screen (n=122) (Figure 1B). These samples were again screened for reactivity by ELISA using POWV EDIII-MBP antigen. A more stringent cut-off of 5 standard deviations (SD) above the mean negative control sera signal (OD 450nm >0.302) was used to determine putative positive samples. Samples meeting this more stringent cut off (n=68) were again screened for reactivity using the ELISA assay but against POWV EDIII lacking MBP, to rule out background binding of sera to MBP (Figure 1C). 14 (1.3%) putative positive samples had a signal 4 SD above the mean negative control sera (OD 450nm >0.447) (Figure 1D). This threshold was chosen based on prior literature and the resulting putative positive samples were further characterized^11^. To mitigate against potential false negative results, a random sample of 60 sera that were negative in ELISA with POWV EDIII-MBP were again screened using POWV-EDIII, and no new positive samples were detected (data not shown). All putative positive samples were from male participants aged 22-24 (Supplementary Table 2). The home state of these individual participants is not known.

**Figure 1:**
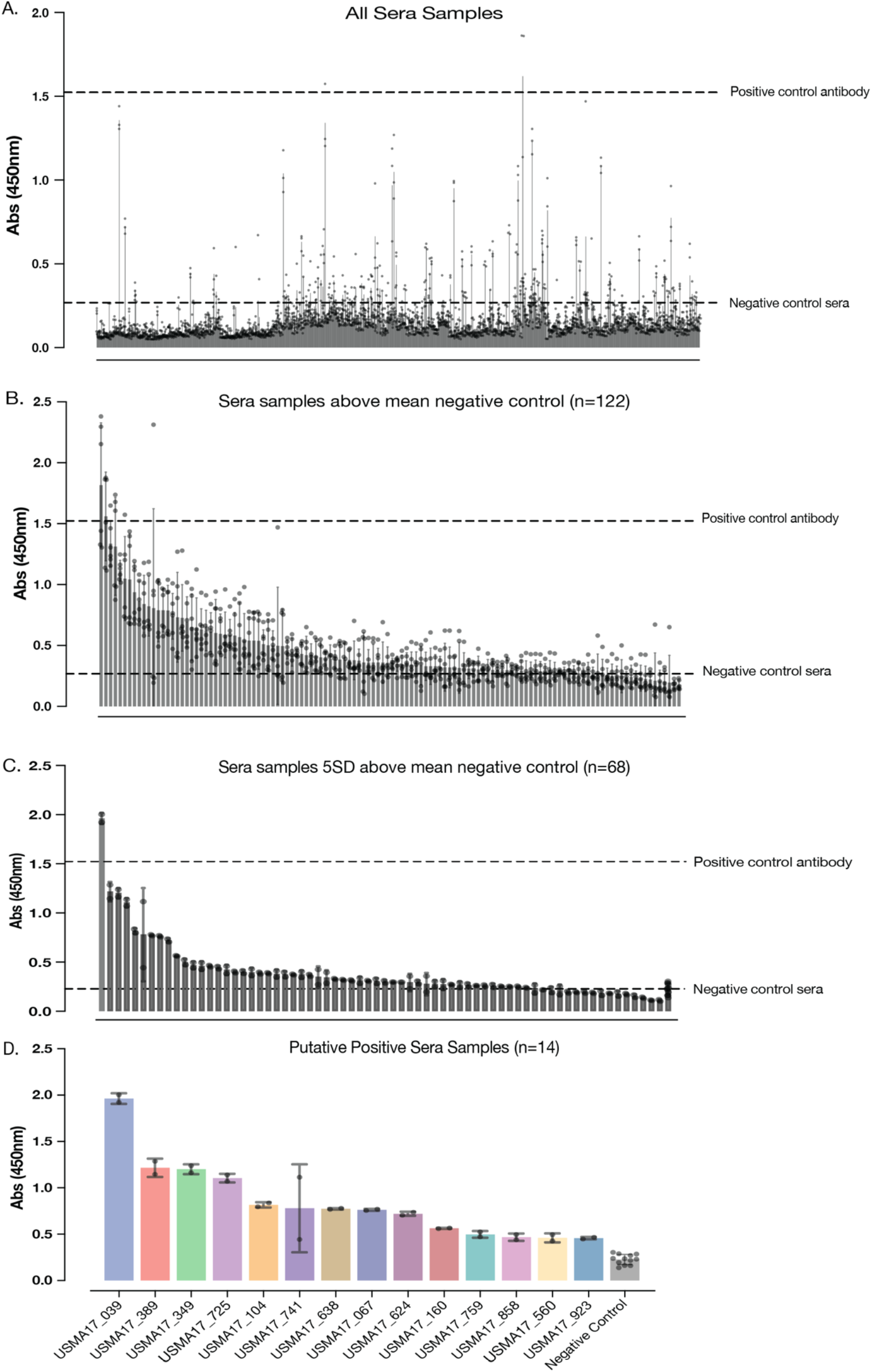
Tiered screening of serum for reactivity to POWV EDIII. Absorbance at 450 nm used in A-D. Positive and negative control thresholds are indicated in A-C. A.) 122/1,051 Cadet “Post” samples were determined to be above mean negative control Abs 450 nm. B.) 122 samples were re-screened and 68 were determined to at least 5 standard deviations above mean negative control Abs 450nm. C.) 68 samples from B. were re-screened using POWV EDIII without MBP tag and 14 samples were 4 standard deviations above the mean negative control Abs 450nm. D.) 14 putative positive samples after multiple tiered screening shown.

### A majority of putative positive samples show evidence of seroconversion during study period

To address the question of whether POWV seropositivity developed during the study period, while the Cadet was enrolled at USMA, the matching “Pre” samples of the 14 positive “Post” samples were analyzed by ELISA with POWV EDIII antigen (Figure 2). Sample USMA17-039 showed the highest signal in both “Pre” and “Post” samples, without a significant difference between the two time points, suggesting this participant entered USMA with prior POWV exposure. 11 of 14 samples showed a statistically significant increase in ELISA signal when the “Post” sample was compared to the “Pre”. These findings suggest that Cadets may have been exposed to POWV and subsequently seroconverted during their time at USMA.

**Figure 2:**
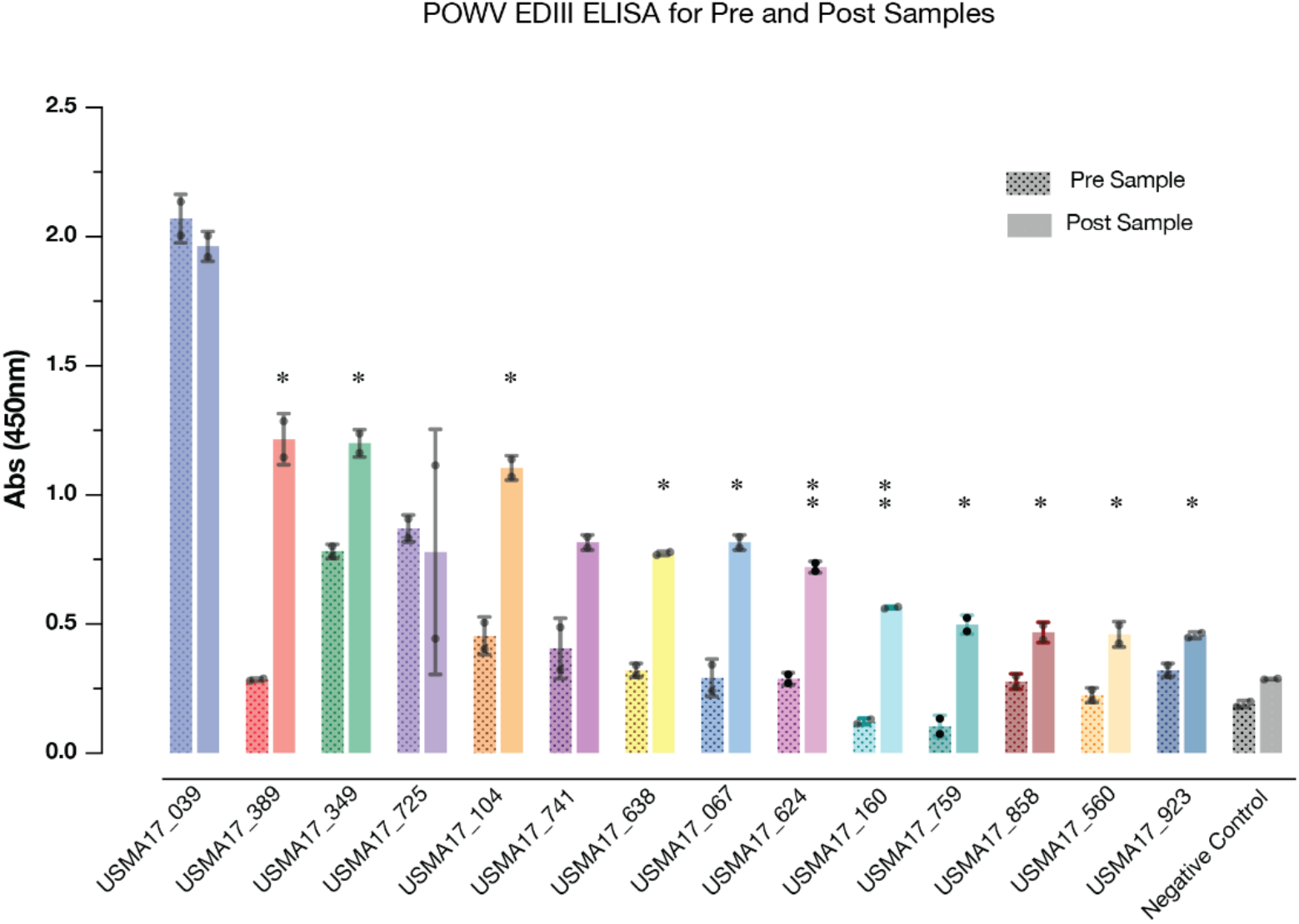
Comparison of POWV EDIII binding in matching “Pre” and “Post” Serum Samples. Pre sample, left, hatched bars and Post sample, right, solid bars, binding to POWV EDIII was compared using ELISA. Mean absorbance from two biological replicants done in triplicate for each paired Pre and Post samples were compared using Welsch’s t test. *p=0.0332, ** p=0.0021. *** p=0.0002, **** p<0.0001.

### Putative positive samples are reactive with POWV reporter virus particles (RVPs)

Prior studies on POWV and other flaviviruses have shown that EDIII is highly immunogenic and an important target for antibodies, making it an ideal screening target for seropositivity^13–15^. To test the reactivity of putative positive sera samples with full-length POWV E in the context of a virion, due to limited availability of samples we tested the 10 samples with the highest signal in the POWV EDIII screen for reactivity against POWV RVPs using an ELISA assay (Figure 3A). All samples tested had a strong signal, suggesting reactivity with POWV RVPs in a similar fashion to POWV EDIII. m158.25, a POWV monoclonal antibody that binds an epitope in EDIII, was used as a positive control and showed a similar signal to positive sera samples^15^.

**Figure 3:**
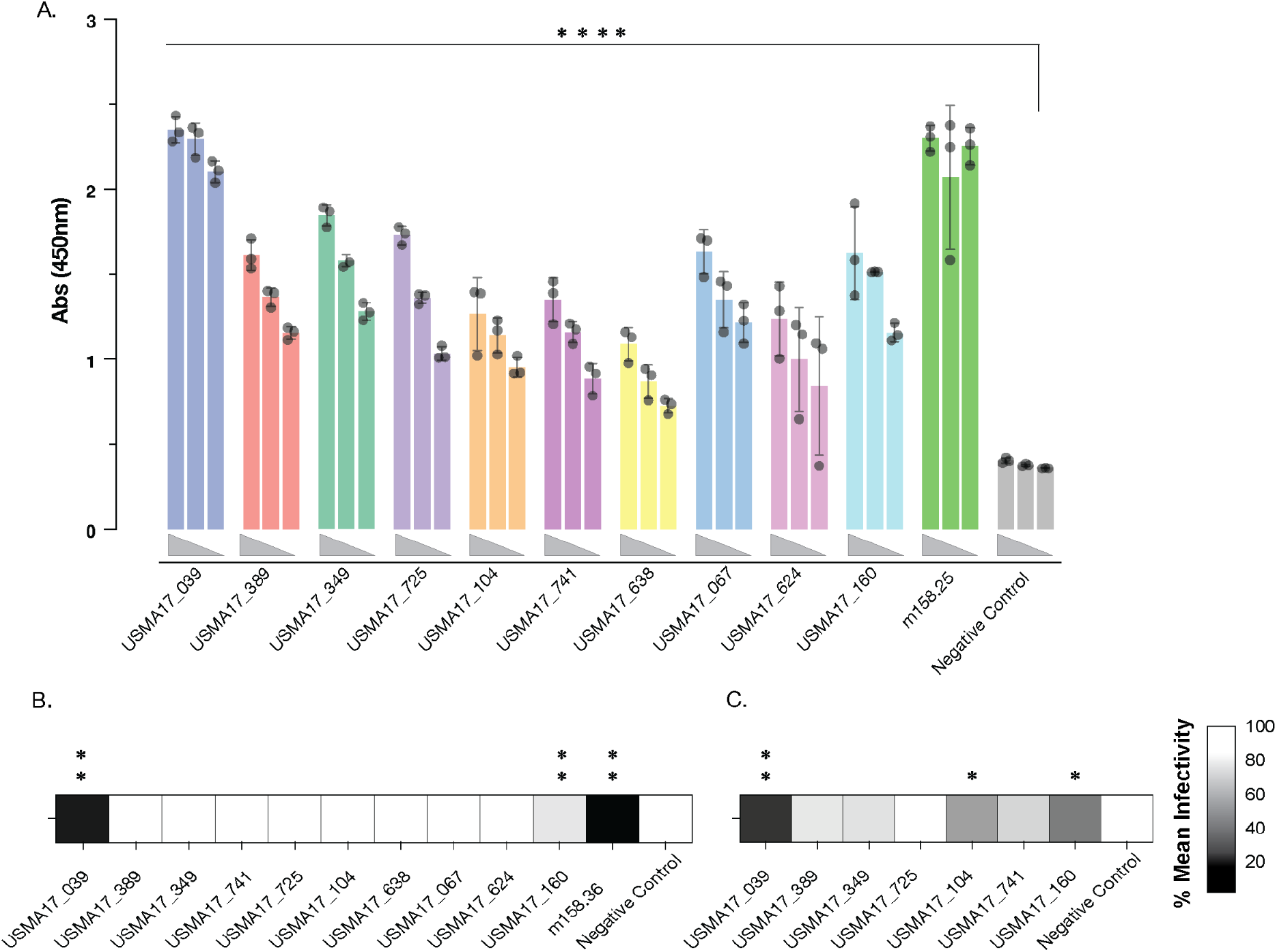
Analysis of putative positive sera samples for reactivity with reporter virus particles (RVP) bearing POWV envelope and neutralization of authentic POWV. A.) POWV RVP ELISA using 3-fold serial dilution series of select positive samples. Mouse POWV mAB m158.25 was included as positive control. Absorbance at 450nm was measured. One biological replicate done in triplicate shown due to limited availability of samples. Samples compared using 2-way ANOVA, significant differences compared to negative control shown. **** p<0.0001. B.) POWV RVP Neutralization assay. Serially diluted sera were incubated with POWV II E RVPs and then added in triplicate to Vero cells. After 24 hours cells were fixed and stained using anti-GFP antibody and infectivity was measured using Cytation 5 automated fluorescence microscope. Mean percent infectivity of highest serum dilution was compared to negative control using Welch’s t test. *p=0.0332, ** p=0.0021. C.) Authentic POWV neutralization. POWV II (Spooner) were incubated with serial dilutions of sera for 1 hour and then added to A549 cells. 48 hours post-infection cells were fixed and stained with anti-NS1 mAb and infectivity was measured using Cytation 5 automated fluorescence microscope. Mean percent infectivity of highest serum dilution was compared to negative control using Welch’s t test. *p=0.0332, ** p=0.0021.

### Sera demonstrating neutralization of POWV RVPs and authentic POWV

The ability of these samples to neutralize POWV RVPs bearing the full E protein was also investigated. Samples USMA17-039 and USMA17-160 showed significant neutralization compared to negative control sera (Figure 3B). To further investigate the neutralization results seen with POWV RVPs, neutralization of authentic POWV was also assessed. The top 7 “post” sera samples with the highest reactivity in POWV EDIII ELISA were selected to evaluate for their ability to neutralize authentic POWV. Again, sample USMA17-039 and USMA17-160 as well as USMA17-104 showed significant neutralization as compared to control sera (Figure 3C).

### Three putative POWV positive samples also show cross-reactivity to West Nile virus EDIII

To examine the cross-reactivity of putative positive samples with another flavivirus, we selected West Nile Virus (WNV) to test as this is a mosquito-borne flavivirus found in the American Northeast, including New York State^20^. ELISA assays were performed using WNV EDIII (Figure 4), with the WNV EDIII-specific monoclonal antibody E16 as a positive control. The majority of putative positive POWV sera samples did not bind WNV EDIII, including USMA17-039, the highest-reactivity sample in the POWV EDIII ELISA, suggesting the POWV EDIII ELISA is largely type-specific, with little or no flavivirus cross-reactivity. However, Cadets could conceivably be exposed to other flaviviruses. Three of the putative positive POWV sera samples also showed statistically reactivity with WNV EDIII when compared to negative control sera (Figure 4), suggesting that the Cadet population at USMA is at risk for exposure to multiple endemic flaviviruses.

**Figure 4.**
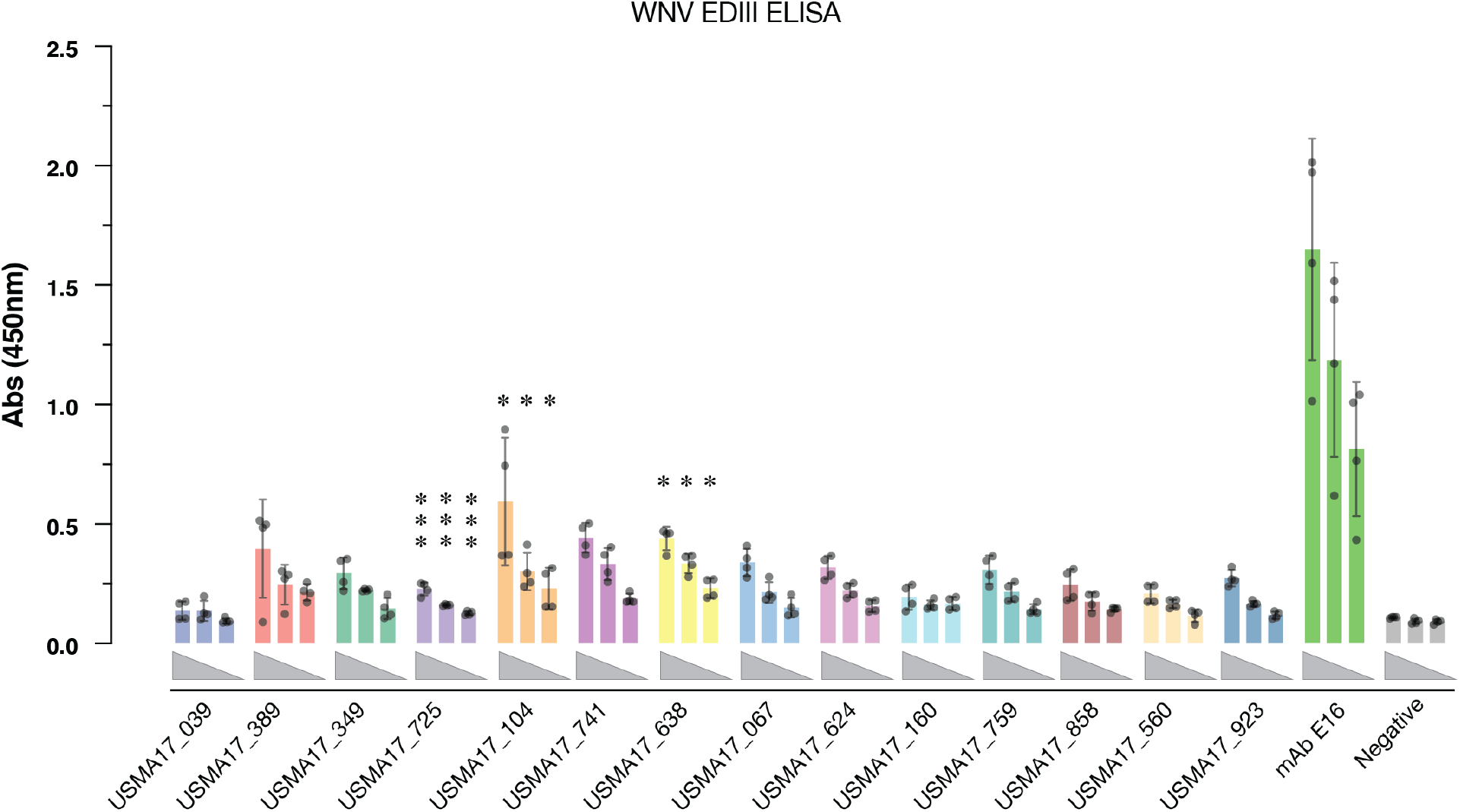
Reactivity of putative positive sera samples with WNV EDIII. WNV EDII ELISA reactivity was determined with three-fold serial dilutions of POWV putative positive sera samples. WNV mAb E16 was included as positive control. Absorbance at 450nm was measured. Data from 2 biological replicates done in duplicate shown. Samples compared using 2-way ANOVA, significant differences compared to negative control shown. *p=0.0332, ** p=0.0021, *** p=0.0002.

### POWV seroprevalence among cadets with a history of Lyme Disease

Limited clinical information was available for de-identified serum samples. Amongst 1,051 “Post” sera, three references to Lyme Disease were annotated in Clinical Comments (Supplementary Table 3). One of these samples, USMA17-741, was among the POWV putative positive samples (Figure 1D) while the other two, USMA17-174 and USMA17-893, were not positive for POWV. Lyme serology of these two samples and the 14 POWV putative positive samples confirmed detection of *B. burgdorferi* antibodies for only USMA17-174 (Supplemental Figure 1). No other POWV putative positive sample was seropositive for Lyme.

## Discussion

Prior seroprevalence studies of POWV report wide ranges of positivity from <1% to 22.9% depending on the population studied^16, 21–23^. These studies, conducted primarily in regions where POWV cases have been reported, including the US Northeast and Upper Midwest, and parts of Canada, include heterogeneous populations ranging from healthy individuals to those with prior tick-borne disease or those living in areas with recent neuroinvasive cases. This variability, along with differences in cohort size, likely contributes to the broad range of reported seroprevalence estimates. In this study, we examined POWV seroprevalence in one of the largest retrospective cohorts (n=1051) described and found a positivity rate of about 1%. In comparison, a cohort study examining a population (n=273) in an area with a recent cluster of neuroinvasive POWV cases (n=3), found a lower POWV positivity rate of <1%^21^. The higher seroprevalence (1.3%) observed in our study may reflect increased tick exposure among cadets, who live in an area with a high burden of *Ixodes* scapularis ticks and engage in frequent outdoor activity during peak tick season. However, the positivity rate in our study is significantly lower than that found in a study of patients in the Northeastern United States with a self-reported history of Lyme Disease, which is also carried by the *Ixodes scapularis* tick (22.9% positivity in EDIII-based assay, n=538)^16^. These differences underscore the need for large cohort studies in at-risk populations to better define POWV exposure in North America.

A key strength of our cohort, in addition to its size and risk factors for tick-borne diseases, is the availability of pre- and post-matriculation samples. This unique feature, combined with the DODSR’s continued storage of samples from subsequent USMA classes, allows assessment of seroconversion over time and comparison of seroprevalence across multiple Cadet cohorts. This could be useful to monitor changes in POWV prevalence in an endemic area and provide a justification for enhanced testing of symptomatic patients if rates are found to be rising significantly over time. Similar assays can also be performed in this cohort to screen for other arboviruses of concern, such as WNV and Eastern equine encephalitis virus (EEEV), both of which have been reported in areas surrounding USMA^24, 25^. In our limited screen, we identified 3 participants who may have also been exposed to WNV (Figure 4), suggesting this cohort may provide significant information about the prevalence of several important arboviruses.

One limitation of this study is the retrospective nature of the evaluation. We are unable to obtain direct evidence of active POWV infection (e.g. positive PCR) positivity at the time of infection and, therefore, must rely on retrospective antibody testing to detect cases. The signal in our assays may be affected by the age of the samples and the length of time between exposure to POWV and when the sample was obtained. For participants with apparent seroconversion, exposure may have occurred outside the USMA region. Further, follow-up sampling was not possible, limiting further characterization of immune responses. These limitations underscore the need for prospective studies with serial sampling to capture real-time seroconversion, track antibody responses, and enable downstream analyses, such as peripheral blood mononuclear cells (PBMCs) antibody isolation.

Severe neuroinvasive POWV disease is a morbid condition with no approved treatments or vaccines. The majority of POWV cases diagnosed in North America are those who present to care with severe disease. Given the paucity of cases, diagnosis of POWV can be challenging and may delay treatment with future approved therapeutics, including antibody-based therapeutics, which are often most effective when administered early in the course of infection^26, 27^. An understanding of the prevalence of non-neuroinvasive infection is essential to improve POWV diagnosis and the development and deployment of POWV-targeted therapeutics.

## Data Availability

All data produced in the present study are available upon reasonable request to the authors

## Author contributions

Conceptualization: SLB, JRL, KC, SLS, KJO, JRH, EHM

Methodology: ALT, ZRD, GF, KEE, GCH, MJR, WVC, MND, KAK, JEJ, AJ, JSC, ARW, DG, SLB, JRH, EHM

Investigation: GF, JRL, ALT, EHM, JEH, GCH, WVC, DG, JRH, EC, CF

Data analysis: BAB, SLB, GL, EHM

Supervision: JRL, KC, CF, ASH, KJO, JRH, EHM

Manuscript preparation: ALT, ZRD, JEH, DG, JRH, EHM

Manuscript editing: ALT, ZRD, JEH, GF, ASH, CF, JRL, KC, KJO, JRH, EHM

We thank Matt Ramirez, Karyme Paez, Narisa Lee for lab management and technical support. We thank K. Dempsey and C. O’Brien for laboratory management; K. Cogliano for programmatic and administrative support. As well as Dr. Joseph R. Loverde for expert technical assistance and Karen Y. Peck in her role as the USMA Human Protections Director.

## Disclosures

J.R.L was supported by the National Institutes of Health (NIH; https://www.niaid.nih.gov/) grant U19AI181960. E.H.M. was supported by the Institute for Clinical and Translational Research at Einstein and Montefiore (https://einsteinmed.edu/centers/ictr) (K12TR004411). C.F was supported by Congressionally Directed Medical Research Programs grant HT9425-24-1-0873. A.L.T and G.F. were additionally supported in by NIH training grants T32-GM149364 (Medical Scientist Training Program) and T32-AI08117 (Geographic Medicine and Emerging Infections). The funders had no role in study design, data collection and analysis, decision to publish, or preparation of the manuscript. The content is solely the responsibility of the authors and does not necessarily represent the official views of the National Institutes of Health.

Opinions, conclusions, interpretations and recommendations are those of the authors and are not necessarily endorsed by the US Department of the Army, the US Department of Defense or the US Department of Health and Human Services.

J.R.L. was a consultant for GlaxoSmithKline and receives funding for unrelated projects from Merck. KC owns equity in Integrum Scientific, LLC and Eitr Biologics, Inc.

---

**Supplemental Figure 1:**
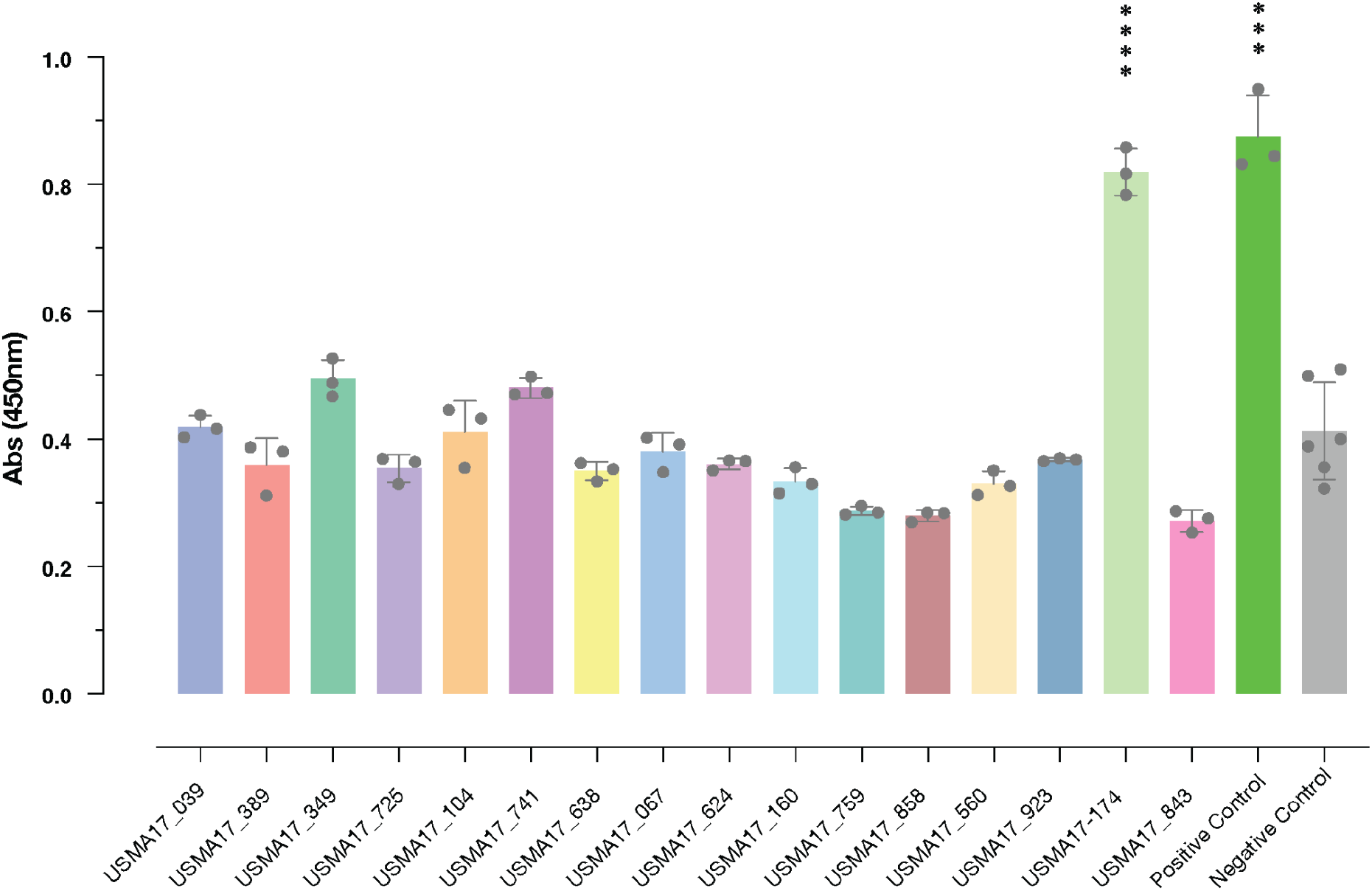
Reactivity of putative positive samples with *Borrelia burgdorferi* antigen. 14 POWV positive samples and two samples known to have tested positive for Lyme Disease were tested for reactivity to *B. burgdorferi* antigen using ELISA assay. One biological replicate with 3 technical replicates shown due to limited sample availability. Samples compared to negative control sera (6 technical replicates) using unpaired t-test with Welch’s correction. *** p=0.0002; **** p<0.0001.

**SUPPLEMENTARY TABLE 1.** HOME STATE OF RECORD FOR 2017 USMA GRADUATING CLASS.

| Region & Division<sup>1</sup> | Total Persons<sup>2</sup> | Percent of total |
| --- | --- | --- |
| **Northeast** | **214** | **22.1** |
| *New England* (CT, ME, MA, NH, RI, VT) | 57 | 5.8 |
| *Middle Atlantic* (NJ, NY, PA) | 157 | 16.2 |
| **Midwest** | **179** | **18.5** |
| *East North Central* (IL, IN, MI, OH, WI) | 132 | 13.6 |
| *West North Central* (IA, KS, MN, MO, NE, ND, SD) | 47 | 4.9 |
| **South** | **365** | **37.7** |
| *South Atlantic* (DE, DC, FL, GA, MD, NC, SC, VA, WV) | 210 | 21.7 |
| *East South Central* (AL, KY, MS, TN) | 52 | 5.4 |
| *West South Central* (AR, LA, OK, TX) | 103 | 10.6 |
| **West** | **205** | **21.2** |
| *Mountain* (AZ, CO, ID, MT, NV, NM, UT, WY) | 70 | 7.2 |
| *Pacific* (AK, CA, HI, OR, WA) | 135 | 13.9 |
| **Island Territories** (Guam, Northern Mariana Islands, Puerto Rico) | **5** | **0.5** |
| **Total<sup>3</sup>** | **968** | **100%** |

<sup>1</sup> Designated by the United States Census Bureau
<sup>2</sup> Numbers correspond to the final 2017 USMA graduating class (n=968), minus students who provided a residence outside the U.S. and/or its territories. The de-identification of patient samples necessitated an inference from the total data (968) for the actual number of samples (938).
<sup>3</sup> Data available for 968 of the 1,051 study participants. 83 participants provided pre-commissioning serum but did not graduate from USMA.

**SUPPLEMENTARY TABLE 2.** POWASSAN VIRUS SEROPREVALANCE.

|  | *Numbers & Percent of Total* | *Gender* | *Race* |
| --- | --- | --- | --- |
| **Positive** | 14/1,051 = 1.3% | 14/14 = 100% M | 11/14 = 78.6% W |
|  |  | 0/14 = 0% F | 2/14 = 14.3% B |
|  |  |  | 1/14 = 7.1% Z |
|  |  |  | 1/56 = 1.8% NA |
| **Negative** | 1,037/1,051 = 98.7% |  |  |

B, black; EDIII, envelope protein domain III; F, female; M, male; NA, not available; O, other; OD, optical density; W, white; Z, unknown

**SUPPLEMENTARY TABLE 3.**
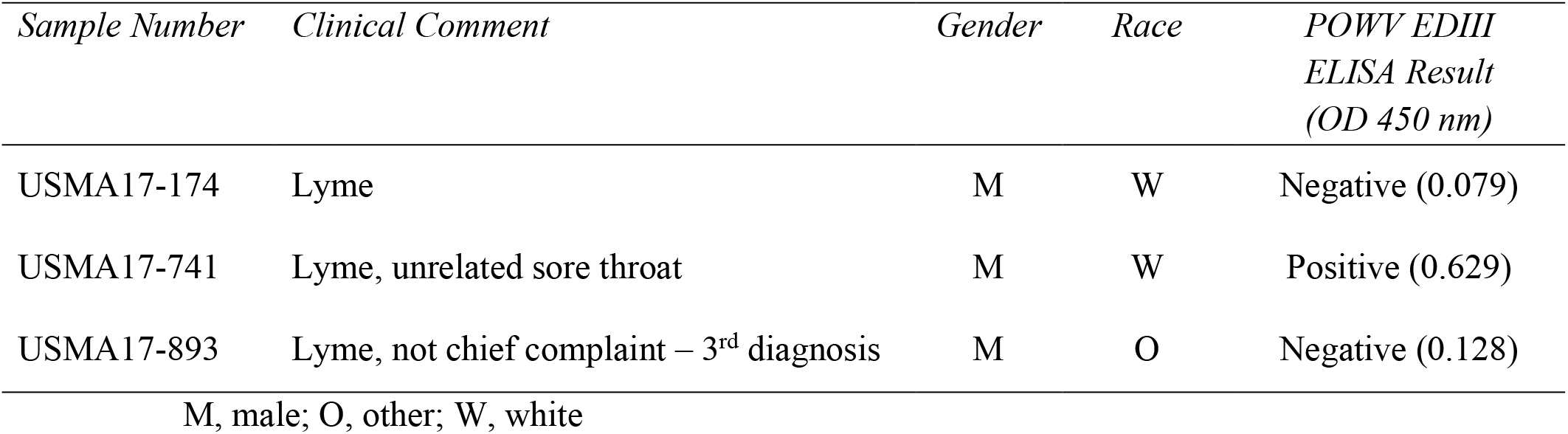
POWV SEROPREVALENCE FOR PARTICIPANTS WITH HISTORY OF LYME DISEASE.

| *Sample Number* | *Clinical Comment* | *Gender* | *Race* | *POWV EDIII*<br>*ELISA Result*<br>*(OD 450 nm)* |
| --- | --- | --- | --- | --- |
| USMA17-174 | Lyme | M | W | Negative (0.079) |
| USMA17-741 | Lyme, unrelated sore throat | M | W | Positive (0.629) |
| USMA17-893 | Lyme, not chief complaint – 3<sup>rd</sup> diagnosis | M | O | Negative (0.128) |

M, male; O, other; W, white

